# Associations Between Body Mass Index, Visual Acuity, and Resting Heart Rate in Adolescents: A Multifactorial Analysis of Physical Examination Data

**DOI:** 10.64898/2026.08.24.26361267

**Authors:** Li Zhiping, Liu Xutao, Teng Xiaopeng, Wang Ping, Zhang Qianqian, Tan Yanhua, Li Haiyang, Zhuang Huaping, Zheng Weibo

**Author notes:** Corresponding authors: Zhuang Huaping, Zheng Weibo. These authors contributed equally to this work (co-first authors) (Li Zhiping, Liu Xutao).

## Abstract

Body mass index (BMI), visual function and resting heart rate (RHR) are standard measurements in adolescent physical examinations, yet their interconnections and age- and gender-specific differences are not fully clarified. This multicenter retrospective cohort study analyzed 130,832 screening records of 8-17-year-old adolescents from 2022 to 2024, using mixed-effects models and stratified analyses to examine BMI’s independent correlations with UCVA (UCVA) and RHR, alongside the moderating effects of age and sex. Boys presented higher BMI values and greater overweight/obesity prevalence, whereas girls had poorer UCVA. After adjusting for confounders, every 1 kg/m² increment in BMI correlated with a −0.008 log MAR change (a stronger effect in girls) and a 0.26 beat-per-minute rise in RHR. The inverse BMI-UCVA correlation peaked at ages 8-11, weakened among 12-16-year-olds, and reversed at age 17. RHR decreased steadily with age, being marginally lower in boys, with a notable age-sex interaction. Age and sex jointly shaped the correlations among the three indicators, whose statistically significant links varied across developmental periods. Therefore, integrated screening should assess all indicators comprehensively while accounting for age and gender disparities. We propose unified, sex and age-stratified adolescent screening and intervention strategies to simultaneously mitigate obesity, myopia and cardiovascular risks, rather than isolated single-disease prevention.

## 1. Background

Over the past decade, the primary health burdens among adolescents have shifted alongside socioeconomic and lifestyle changes, with obesity recognized as one of the most serious public health challenges of the 21st century^[1]^. From 2010 to 2019, the prevalence of overweight/obesity among primary and secondary school students in China increased by 8.7 percentage points, with a sustained upward trend across urban and rural settings and all age groups^[2]^. Evidence indicates that approximately 70% of adolescents with obesity have at least one cardiovascular risk factor, and more than 25% present with two or more concurrent risk factors^[3]^. Adolescent obesity is also a significant predictor of all-cause mortality (ACM) in adulthood^[4]^.

Myopia is particularly prevalent in East and Southeast Asia. A 2020 report from China estimated the overall prevalence of myopia among children and adolescents at 52.7%^[5]^, with increasingly earlier onset^[6]^; projections suggest that by 2050, the prevalence may approach 84%^[7]^. Apart from genetic predisposition, myopia has been associated with eye exercises^[8]^, time spent outdoors^[9]^, sleep patterns and academic workload^[10, 11]^, visual/dietary habits and parental guidance^[12]^, as well as environmental exposures (e.g., suboptimal study lighting, PM2.5/nitrogen oxides)^[13, 14]^. Several studies have demonstrated a positive association between higher BMI and myopia risk^[15, 16]^. Overweight and obesity are also independent risk factors for adolescent hypertension and other cardiovascular conditions^[17–20]^; elevated intraocular pressure has been linked to myopia in some studies^[21, 22]^, suggesting that blood pressure/intraocular pressure may partially mediate the relationship between adiposity and refractive risk.

Despite growing attention to the interrelations among key physiological indicators in adolescence-body mass index (BMI), vision, and heart rate ^[23, 24]^—three critical gaps remain. First, systematic integrative evidence quantifying the independent associations between BMI and both visual acuity and heart rate within the same adolescent cohort is limited ^[25, 26]^. Second, heterogeneity across critical modifiers such as sex and age has not been adequately characterized ^[27]^, constraining the development of precision public health strategies. Third, existing prevention and control efforts tend to target single problems (obesity, myopia, or cardiovascular risk) in isolation, despite evidence suggesting shared underlying mechanisms and modifiable risk factors ^[28, 29]^. To address these gaps, the present study uses routine health screening data from 130,832 examination records of adolescents aged 8-17 years (2022-2024) to address the following research questions: (1) What are the independent associations of BMI with UCVA and resting heart rate, after adjusting for age, sex, and other covariates; (2) Do these associations differ by sex and age; (3) What are the implications of these findings for integrated adolescent health screening strategies.

## 2. Methods

### 2.1 Study Design

We conducted a multicenter retrospective cohort study with dynamic tracking based on consecutive health examination data from 2022 to 2024. Both multiple-group and longitudinal association analyses were performed in adolescents aged 8-17 years. Annual data were integrated to examine sex-age interactions and temporal trends in BMI, visual acuity, and heart rate, with stratified analyses and covariate adjustment to explore developmental patterns. The study included a total of 130,832 examination records from 68,543 male and 62,289 female adolescents aged 8-17 years (2022-2024). Because some participants contributed multiple annual records, each examination was treated as a repeated measurement. Mixed-effects models with random participant intercepts were used to account for within-subject correlation, and sensitivity analyses restricted to one randomly selected examination per participant were performed to verify robustness of the findings.

### 2.2 Data Sources

Data were extracted from annual school-based health screening records (2022-2024) in the study area. The core indicators included: body mass index (BMI, kg/m²), uncorrected visual acuity (UCVA, decimal chart, log-transformed to log MAR for analysis), and resting heart rate (RHR, bpm). Covariates included age, sex, height, weight, and examination year. Detailed measurement protocols and variable definitions are provided in Section 2.4. The dataset used in this study was formally accessed and retrieved for research analysis on 06/01/2026.

### 2.3 Inclusion and Exclusion Criteria

Inclusion: (i) age 8-17 years; (ii) ≥ 1 complete examination (BMI, visual acuity, heart rate, and key covariates); (iii) permanent residence in the study area (≥ 6 months/year).

Exclusion: (i) severe organic disease (e.g., congenital heart disease, diabetes mellitus, thyroid dysfunction, or organic ocular pathology); (ii) missing critical indicators (≥ 2 items) or absence from examinations for three consecutive years; (iii) special physiological states (e.g., pregnancy) or use of medications affecting metabolism or cardiovascular function.

### 2.4 Definitions and Measurements

BMI classification: We applied two complementary systems. (1) WHO age- and sex-standardized Z-scores: underweight (Z < −2), normal weight (−2 ≤ Z ≤ 1), overweight (1 < Z ≤ 2), and obesity (Z > 2). (2) Chinese national standard WS/T 586-2018 using percentiles from national reference data: underweight (< P5), normal weight (P5-P85), overweight (P85-P95), and obesity (≥ P95). The WS/T 586-2018 system was used as the primary classification for all main analyses, while WHO Z-scores were applied in sensitivity analyses to assess consistency across classification standards.

**Height and weight:** measured with an electronic stadiometer/scale (RGZ-160; accuracy ±0.1 kg/±0.1 cm) calibrated daily; extreme values were logically validated (BMI > 40 or < 10 flagged for review).

**Visual acuity:** measured using a standard logarithmic chart under 800 ± 50 lux illumination; recorded using the standard decimal chart (range 0.1-5.0). For regression analyses, decimal acuity was logarithmically transformed to log MAR (logarithm of the minimum angle of resolution) using the formula: log MAR = log₁₀(1/decimal). In this scale, a lower log MAR value indicates better vision, and 0.0 log MAR corresponds to decimal acuity of 5.0 (20/20 Snellen equivalent). Visual impairment was defined as decimal UCVA ≤ 0.8 (Snellen ≤ 20/25) in either eye. Equipment was calibrated quarterly; examiners were uniformly trained (kappa ≥ 0.85).

**Resting heart rate (RHR):** measured using a Polar H10 sensor (beat-to-beat accuracy ±1 ms), recorded as the average over 1 min after ≥ 5 min of seated rest in a quiet environment. For the full age range (8-17 years), tachycardia was defined as RHR > 100 bpm and bradycardia as RHR < 60 bpm, based on pediatric reference standards. Examinations following vigorous exercise or significant emotional arousal within 2 h were excluded. Heart rate variability (HRV), a measure of the variation in time intervals between consecutive heartbeats that reflects autonomic nervous system regulation of cardiac function, was also extracted from Polar H10 recordings when available (SDNN ≥ 100ms, RMSSD ≥ 30ms) and used as supplementary markers of autonomic function in a subset of participants.

All variables used in the main analyses are summarized in Supplementary Table S1, including their definitions, measurement units, and coding schemes. 2.5 Statistical Analysis

### 2.5 Statistical Analysis

Continuous variables are reported as mean ± standard deviation (SD) or median (interquartile range); categorical variables as frequency (%). Normality was assessed using the Shapiro-Wilk test. Two-group comparisons used Student’s t-test or the Mann-Whitney U test; multiple-group comparisons used ANOVA or Kruskal-Wallis tests. Categorical variables were compared using the χ² test or Fisher’s exact test. Post hoc comparisons applied Bonferroni correction.

For multivariable analyses, interaction terms (e.g., BMI × sex; age × sex) were included in linear/logistic regression models and evaluated by likelihood-ratio or Wald tests (*p* < 0.10 for further stratification).

Statistical significance was set at two-tailed α = 0.05. To control the false discovery rate, the Benjamin-Hochberg procedure was applied (FDR < 5%).

To address potential residual confounding, we constructed a directed acyclic graph, based on prior literature to identify the minimal sufficient adjustment set. In sensitivity analyses, we additionally adjusted for school-level clustering using cluster-robust standard errors and applied propensity-score stratification (quintiles) to assess the robustness of the BMI-vision association. E-values were computed to evaluate how strongly an unmeasured confounder would need to be associated with both exposure and outcome to fully explain the observed associations.

### 2.6 Ethics statement

This study was conducted in accordance with the Declaration of Helsinki. The study protocol was reviewed and approved by the Ethics Committee of Zhoupu Hospital, Pudong New Area, Shanghai (Approval No. 2025-C-246). Written informed consent was obtained from all adolescent participants and their parents or legal guardians prior to data collection.

## 3. Results

### 3.1 Cohort Characteristics

The analysis included 130,832 examination records from adolescents aged 8-17 years; 52.4% were male and 47.6% were female. Mean age was 11.5 ± 2.5 years. Normal weight, overweight, and obesity accounted for 68.7% (n = 89,882), 17.2% (n = 22,503), and 14.1% (n = 18,447) of records, respectively. BMI distributions were higher among boys than girls (p < 0.001). Reduced UCVA was present in 64.3% of records and was more frequent among girls than boys (68.9% vs. 60.1%, p < 0.001). RHR ranged from 45 to 129 beats/min; age- and sex-specific summaries are reported in Section 3.5. Examination records were relatively evenly distributed from ages 8 to 13, decreased at ages 14-15, and were sparse at ages 16-17 (Figure 1); estimates for late adolescence should therefore be interpreted with greater caution.

**Figure 1.**
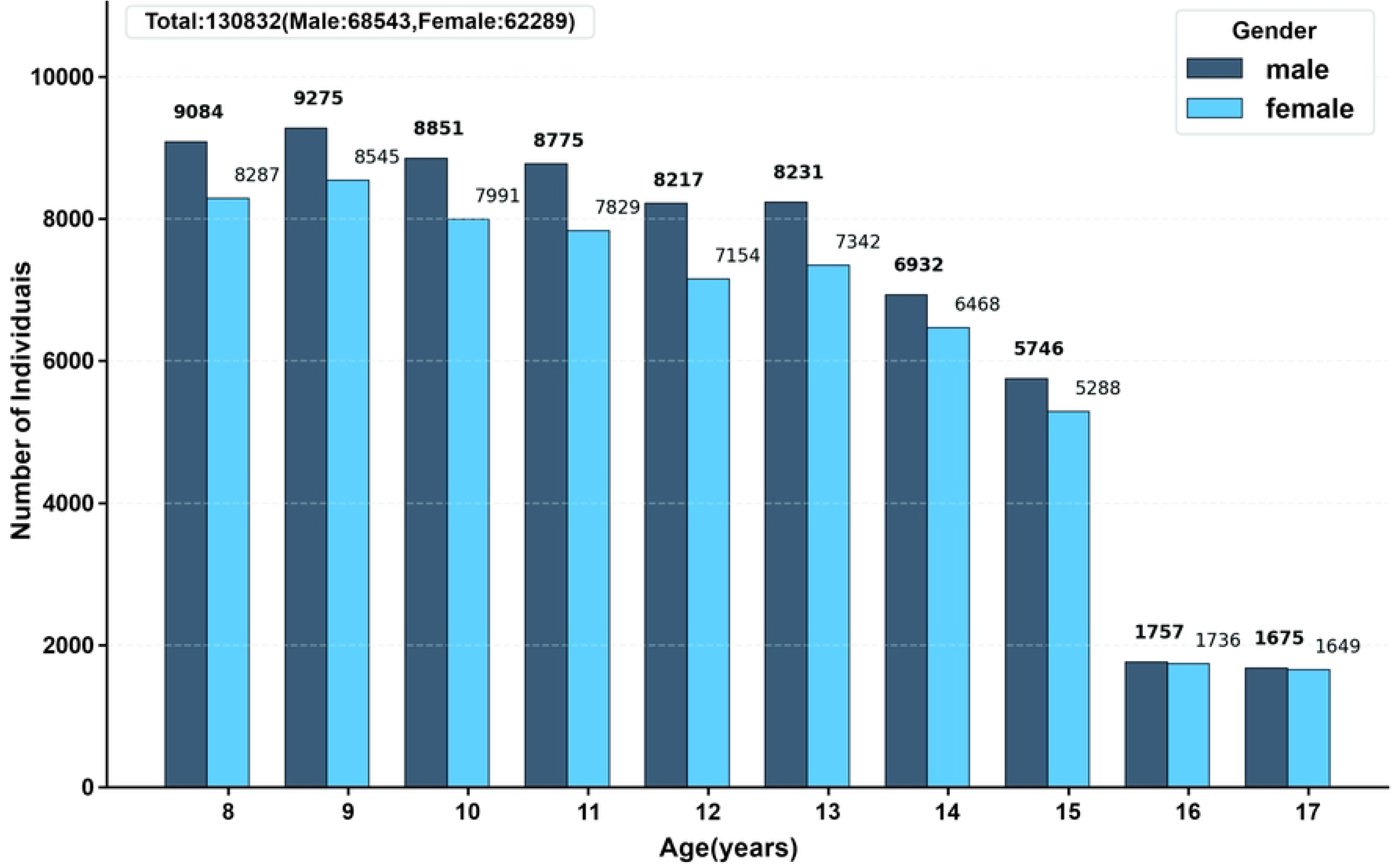
Age- and gender-based sample size distribution (8-17 years). Note: This bar chart displays the number of male and female individuals across ages 8 to 17 years. The total sample size is 130,832, consisting of 68,543 males and 62,289 females. For each age group, the height of the bars corresponds to the count of individuals in that gender category.

In adjusted models, each 1-kg/m² increase in BMI was associated with a −0.008-unit change in UCVA log MAR (95% CI, −0.009 to −0.007; p < 0.001). The BMI-by-sex interaction was significant (p = 0.008), with coefficients of −0.011 (95% CI, −0.013 to −0.009) in girls and −0.005 (95% CI, −0.007 to −0.003) in boys. Each 1-kg/m² increase in BMI was also associated with a 0.26-beats/min increase in RHR (95% CI, 0.21 to 0.31; p < 0.001), although model fit was modest (R² = 0.033).

### 3.2 Analysis of the Correlation Between BMI and Age-Sex Characteristics

#### 3.2.1 Univariate analysis of age on BMI

One-way ANOVA revealed a significant main effect of age on BMI (F = 38,149.49, *p* < 0.001), indicating systematic age-related variation in adolescent BMI. The age-stratified violin plots further demonstrate that, from 8 to 17 years, the BMI distribution gradually shifts upwards and becomes more right-skewed, with increasingly prominent high-BMI tails in mid to late adolescence (Figure 2A). Prior to age 12, observations with BMI ≥ 30 are relatively uncommon for either sex, whereas during puberty and late adolescence (15-17 years) high-BMI cases become markedly more frequent, forming dense upper clusters-particularly among boys. These patterns denote substantial between-age-group heterogeneity, with population differences amplified in mid to late adolescence. Collectively, the results identify age as a key determinant shaping the developmental trajectory of BMI-plausibly reflecting the combined influences of pubertal growth spurts and metabolic adaptations during the transition to adulthood. Practically, they suggest that current adolescent-focused weight interventions may lack sufficient targeting and intensity. Our results suggest that adolescence represents a ‘critical window’ for weight management, with priority allocation of public health resources, strengthened age-stratified surveillance systems, and nutrition and physical-activity strategies calibrated to developmental stage.

**Figure 2.**
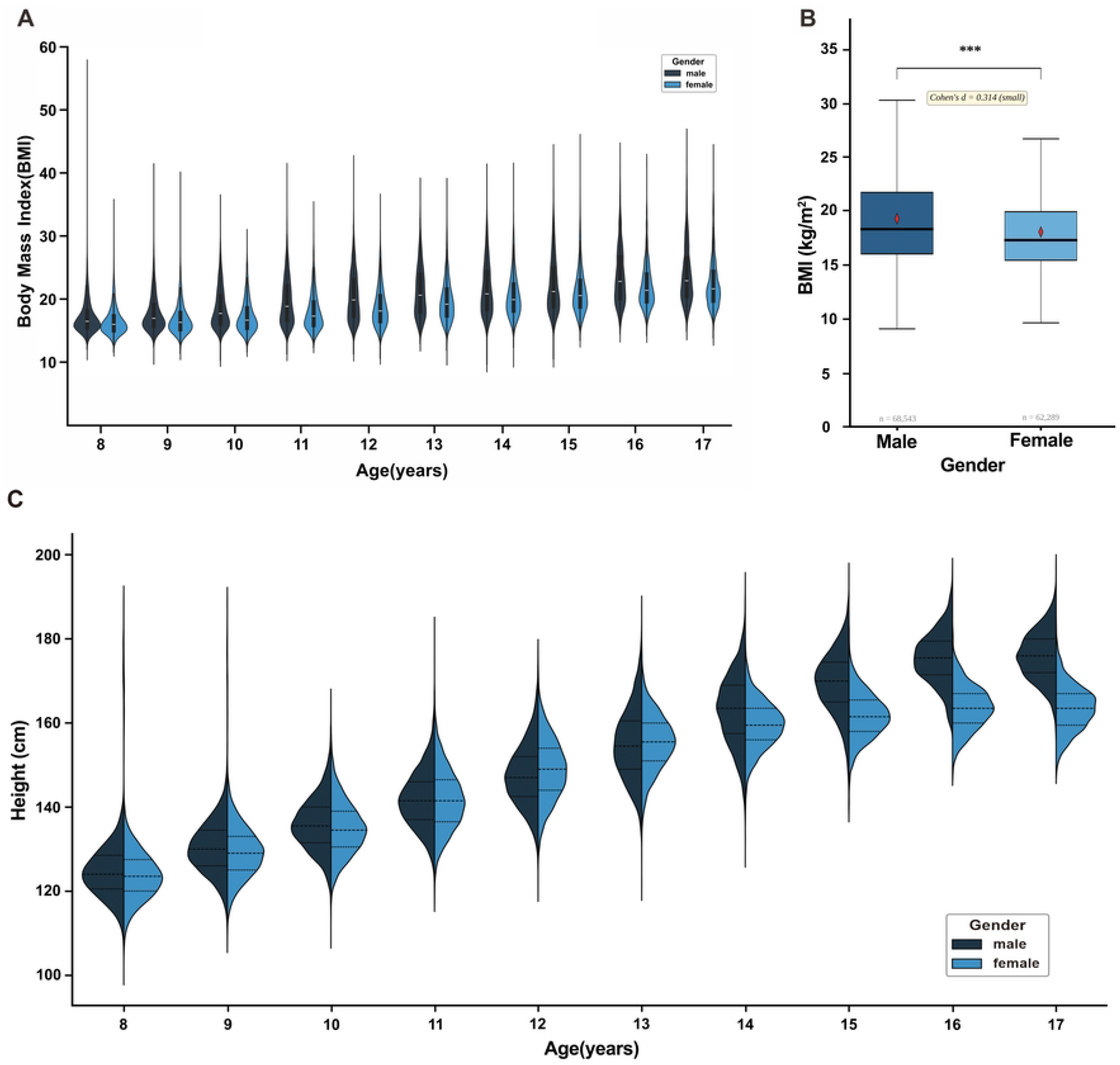
Age- and sex-stratified distributions of BMI and height in adolescents. Note: (A) Violin plots showing the distributions of BMI by age and sex among adolescents aged 8-17 years; the outer contour reflects the kernel density of BMI, the white dot denotes the median, the thick bar shows the 25th-75th percentile range, and the thin line indicates the overall range. (B) The floating bar displays the interquartile range (IQR, Q1-03), the thick horizontal line inside each bar represents the median; the red diamond marks the mean; whiskers extend to the most extreme value within 1.5xIQR. Welch’s t-test: t= 57.253, *p* < 0.001 ***, Cohen’s d= 0.314 (small). (C) Violin plots of height by age and sex. Height increases with age in both sexes; males surpass females from ∼13-14 years, with the gap widening through adolescence and plateauing around age 17.

#### 3.2.2 Univariate analysis of BMI by sex

One-way analysis of variance (ANOVA) showed that sex is a highly significant determinant of BMI (F (1, N) = 4297.20, *p* < 0.001). The boxplots illustrate that boys have a higher median BMI and a broader upper tail than girls, indicating greater dispersion and a larger number of high-BMI outliers among male adolescents (Figure 2B) . Consistent with this distributional pattern, the combined prevalence of overweight and obesity was substantially higher in boys than in girls, and this disparity remained stable across the study period. These findings indicate that boys are at higher risk for overweight and obesity and should be prioritized for tailored, school- and community-based weight-management programs. The persistence of this sex difference also suggests shortfalls in the reach and responsiveness of current health-promotion strategies for boys-particularly with respect to environmental supports and resource provision for key behaviors such as diet, physical activity, sleep duration, and screen time. Accordingly, sex-responsive intervention pathways aligned with the characteristics of male adolescents are warranted.

#### 3.2.3 Height trajectories across adolescence by sex

Height exhibited a pronounced developmental trajectory across the 8-17-year age range, with a total increase of 45.2 cm from 124.6 cm at age 8 to 169.8 cm at age 17. Marked sex differences in growth patterns were confirmed by two-way ANOVA, which revealed a highly significant age × sex interaction (F = 816.16, p < 0.001), indicating that the growth trajectories of male and female adolescents followed fundamentally different patterns (Figure S1A). As illustrated in the violin plot, height distributions demonstrated a progressive rightward shift with age in both sexes, reflecting typical pubertal growth spurts. A notable crossover pattern was observed: at ages 8-11, males were slightly taller than females by approximately 1 cm, but at age 12, females temporarily surpassed males (148.91 vs. 147.58 cm, p < 0.001), reflecting the earlier onset of the pubertal growth spurt in females. Peak height velocity occurred at 11-12 years in females (7.19 cm/yr) and at 13-14 years in males (8.10 cm/yr). The average height of males began to surpass that of females around ages 13-14, with the gap widening progressively through mid-to-late adolescence, reaching 12.42 cm by age 17 (175.99 vs. 163.57 cm; Cohen’s d = 2.23, p < 0.001) (Figure S1B). The full distributions likewise shifted upward with age and showed widening sex differences from approximately ages 13-14 (Figure 2C). By approximately age 17, height distributions stabilized in both sexes, indicating the attainment of adult stature, and the sex difference plateaued. These patterns align with established pubertal timing differences, wherein males typically enter peak height velocity later than females but achieve greater final adult height due to a longer growth period and higher peak velocity. The observed trajectories underscore the importance of considering sex-specific growth patterns when interpreting anthropometric and health indicators in adolescents.

### 3.3 Analysis of the Association Between Body Mass Index and Visual Acuity

#### 3.3.1 Univariate Analysis of Age on Visual Acuity

Regression analysis indicates a significant negative association between age and unaided visual acuity from ages 8-17 (left eye β = −0.0476; right eye β = −0.0527; both p < 0.001), corresponding to an average decline of approximately 0.05 units per year. As shown in the age-specific violin plots comparing left- and right-eye acuity (Figure 3A), the entire distribution shifts leftwards and becomes progressively broader with increasing age, indicating both a growing myopia burden and increasing inter-individual heterogeneity. This pattern highlights the pre-/peri-pubertal period as a critical window for myopia prevention and control, warranting earlier refractive screening, behavioral interventions (reduced near work, increased outdoor time), and risk-stratified, individualized management for high-risk students.

**Figure 3.**
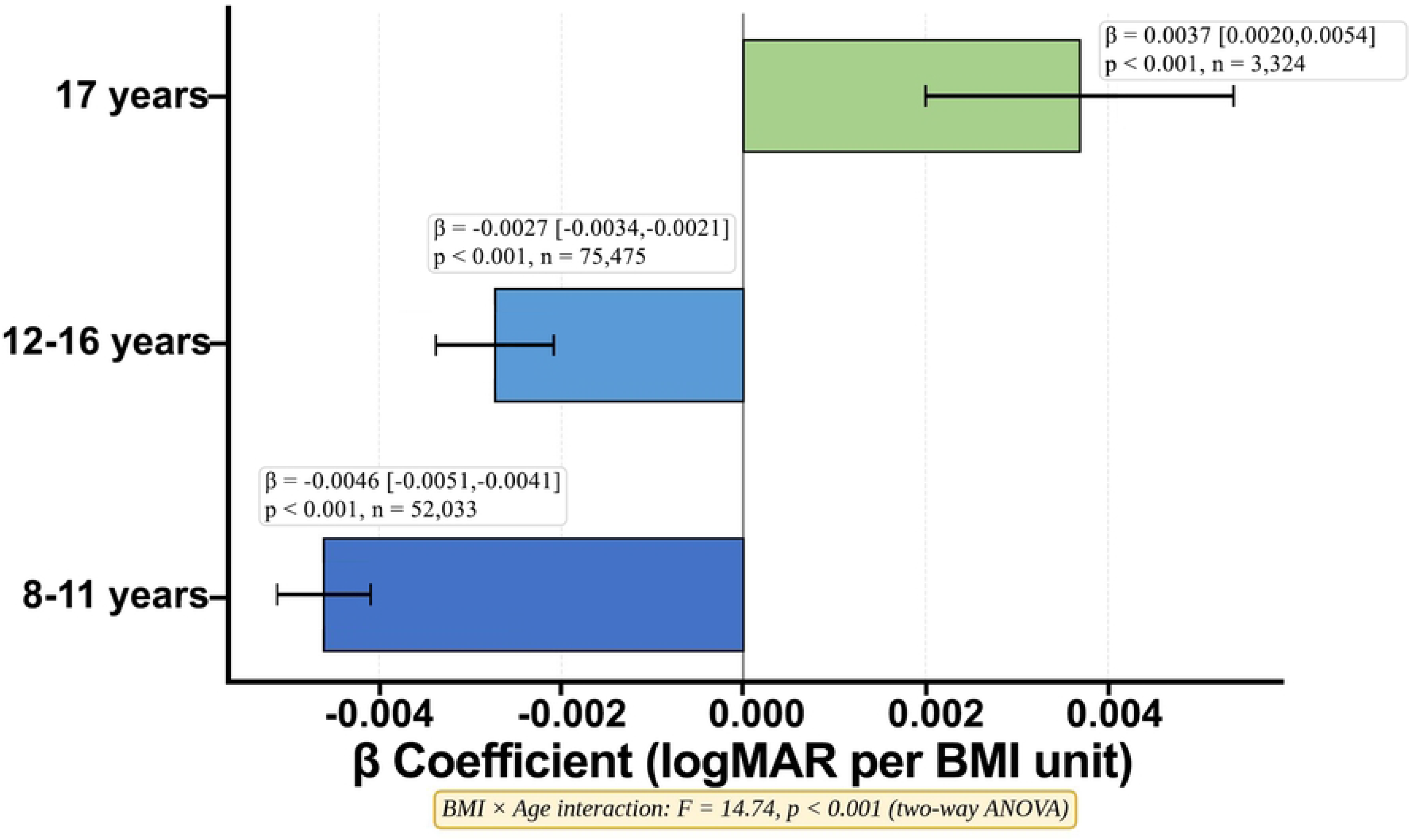
Age- and sex-specific distributions of UCVA among adolescents. Note: (A) Violin plots showing the distributions of UCVA in the left and right eyes across ages 8-17 years; the width of each violin reflects the kernel density of UCVA values at a given age, and the dashed horizontal line denotes the normal-vision threshold (UCVA = 5.0). (B) Boxplots of right-eye UCVA by sex. (C) Boxplots of left-eye UCVA by sex. Boxplots represent interquartile ranges, with the horizontal line inside the box denoting the median, whiskers indicating the overall range, and hollow circles marking outliers.

#### 3.3.2 Univariate Analysis of Sex on Visual Acuity

Significant and consistent sex disparities in UCVA were observed throughout the age range studied, with males consistently exhibiting better average visual acuity than females. The boxplots of left-and right-eye UCVA (Figure 3B and C) clearly show higher medians and slightly narrower lower tails in males compared with females.

Concurrently, increased intra-group variability was evidenced by rising standard deviations (from 2.25 to 4.04 in females and from 2.68 to 4.73 in males), reflecting amplified individual differences post-puberty. These patterns suggest that sex-associated behavioral and biological factors—including near-work demands, outdoor activity exposure, sleep and screen time patterns, and potential endocrine-mediated pathways-exert distinct modulatory effects on visual health. The implementation of sex-sensitive myopia prevention is warranted, entailing earlier and more frequent refractive screening and tailored behavioral interventions for female students and other at-risk populations. Concurrently, fostering supportive school and home environments is essential, with an emphasis on promoting outdoor time, optimizing ergonomic and lighting conditions, and ensuring consistent sleep-wake cycles.

#### 3.3.3 Univariate Analysis of BMI on Visual Acuity

Analysis of descriptive statistics and correlations identified the association between BMI and UCVA as age-dependent. A two-way ANOVA revealed a significant BMI×age interaction on visual acuity (F = 14.74, p < 0.001; Figure 4), confirming that the BMI-vision relationship varied substantially across developmental stages. Age-stratified linear regression further quantified this heterogeneity: in the 8-11-year age group, each 1-unit increase in BMI was associated with a −0.0046 log MAR change in visual acuity (95% CI: −0.0051, −0.0041; p < 0.001); in the 12-16-year group, the association attenuated to β = −0.0027 (95% CI: −0.0034, −0.0021; p < 0.001); and in the 17-year group, the association reversed direction (β = +0.0037, 95% CI: 0.0020, 0.0054; p < 0.001). These results indicate that the BMI-vision association is strongest in early adolescence (8-11 years), progressively weakens through mid-adolescence, and reverses in late adolescence. This phase is also characterized by a marked increase in visual acuity variability, occurring alongside substantial BMI fluctuations, which points to a potential interactive dynamic between these two health indicators. These results strongly indicate that early adolescence constitutes a critical window for integrated health management. Consequently, this calls for the implementation of coordinated screening and intervention protocols that synergistically target weight control and myopia prevention. Priority should be placed on optimizing nutritional intake and physical activity, promoting outdoor exposure, and standardizing near-work habits and sleep schedules.

**Figure 4.**
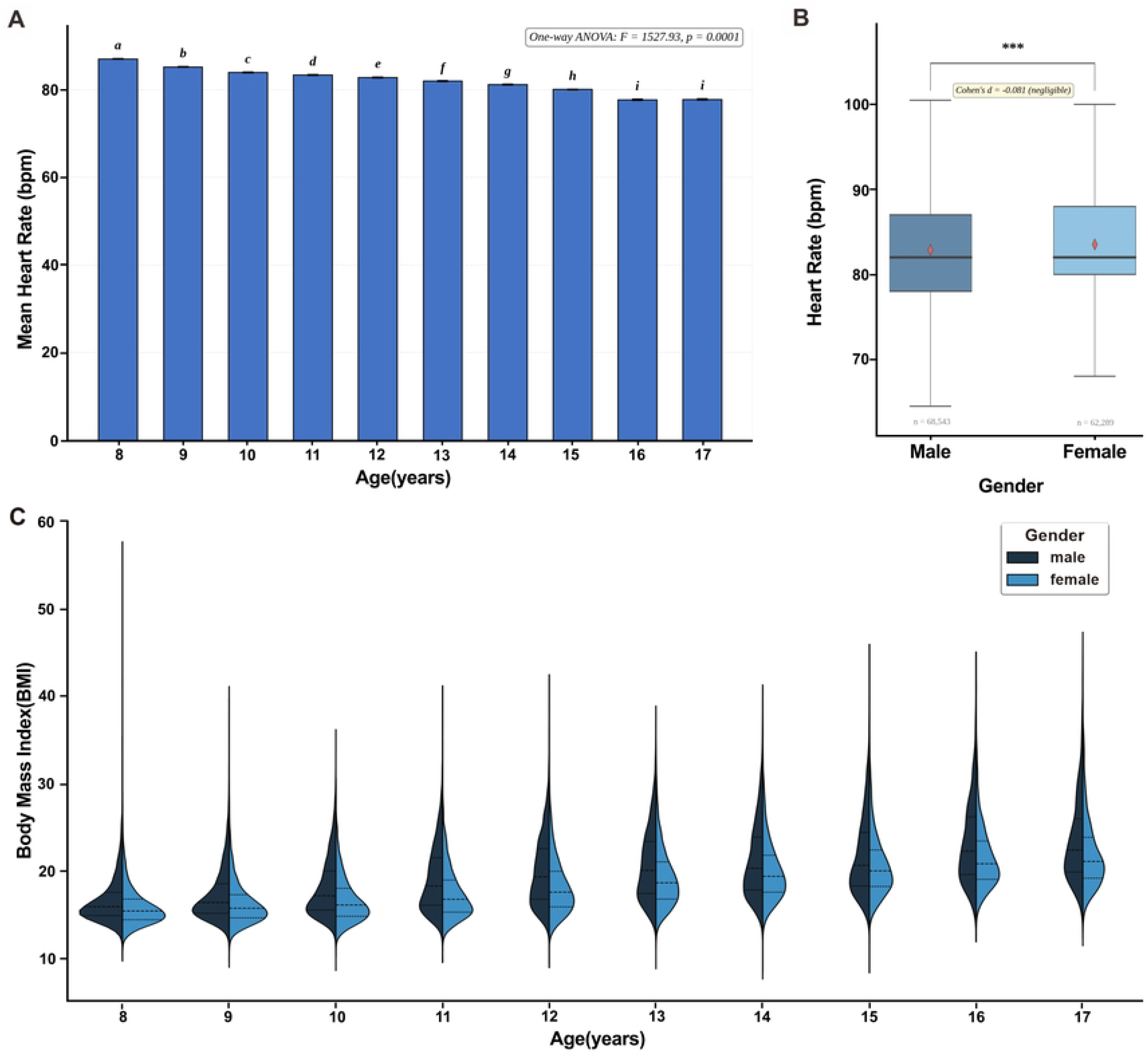
Age-stratified association between BMI and log MAR UCVA. Note. Horizontal bars represent regression β coefficients (per 1 kg/m² BMI increase) for three age subgroups (8-11 years, 12-16 years, 17 years). Horizontal error bars denote 95% CIs; panel labels report β values, 95% CI bounds, p-values, and subgroup sample sizes. The vertical solid line at β = 0 marks the null association threshold. Negative β values indicate higher BMI correlates with lower logMAR (better UCVA); positive β values indicate higher BMI correlates with higher logMAR (worse UCVA). Two-way ANOVA confirmed a significant BMI-by-age interaction (F = 14.74, p < 0.001).

### 3.4 Correlation Analysis of Age and Sex with Heart Rate

#### 3.4.1 Univariate analysis of heart rate by age and sex

Mean resting heart rate showed a pronounced decline across age groups, from 87.1 bpm at age 8 to 77.8 bpm at age 17 (Figure. 5A), corresponding to a mean decrease of approximately 0.94 bpm per year (β = −0.94, 95% CI: −0.96 to −0.93, p < 0.001). A modest but statistically significant sex difference was also observed, with males exhibiting slightly lower mean heart rate than females (82.9 vs. 83.5 bpm; β = −0.64, 95% CI: −0.72 to −0.56, p < 0.001; t-test: t = −14.68, p < 0.001). However, the sex-stratified boxplots (Figure. 5B) revealed largely overlapping distributions, with the magnitude of the sex difference remaining modest (approximately 0.6 bpm) across the age range. The age-related decline in heart rate was monotonic and consistent in both sexes, with a significant age × sex interaction confirmed by two-way ANOVA (F = 12.05, p < 0.001), indicating that the rate of decline differed slightly between males and females. Collectively, while both age and sex were statistically significant predictors of resting heart rate given the large sample size, their effect sizes remained modest in absolute terms. Nevertheless, in clinical practice, adjusting for age and sex and exploring potential interaction terms in cardiovascular monitoring of adolescents remains important to ensure accurate risk stratification and to avoid residual confounding.

**Figure 5.**
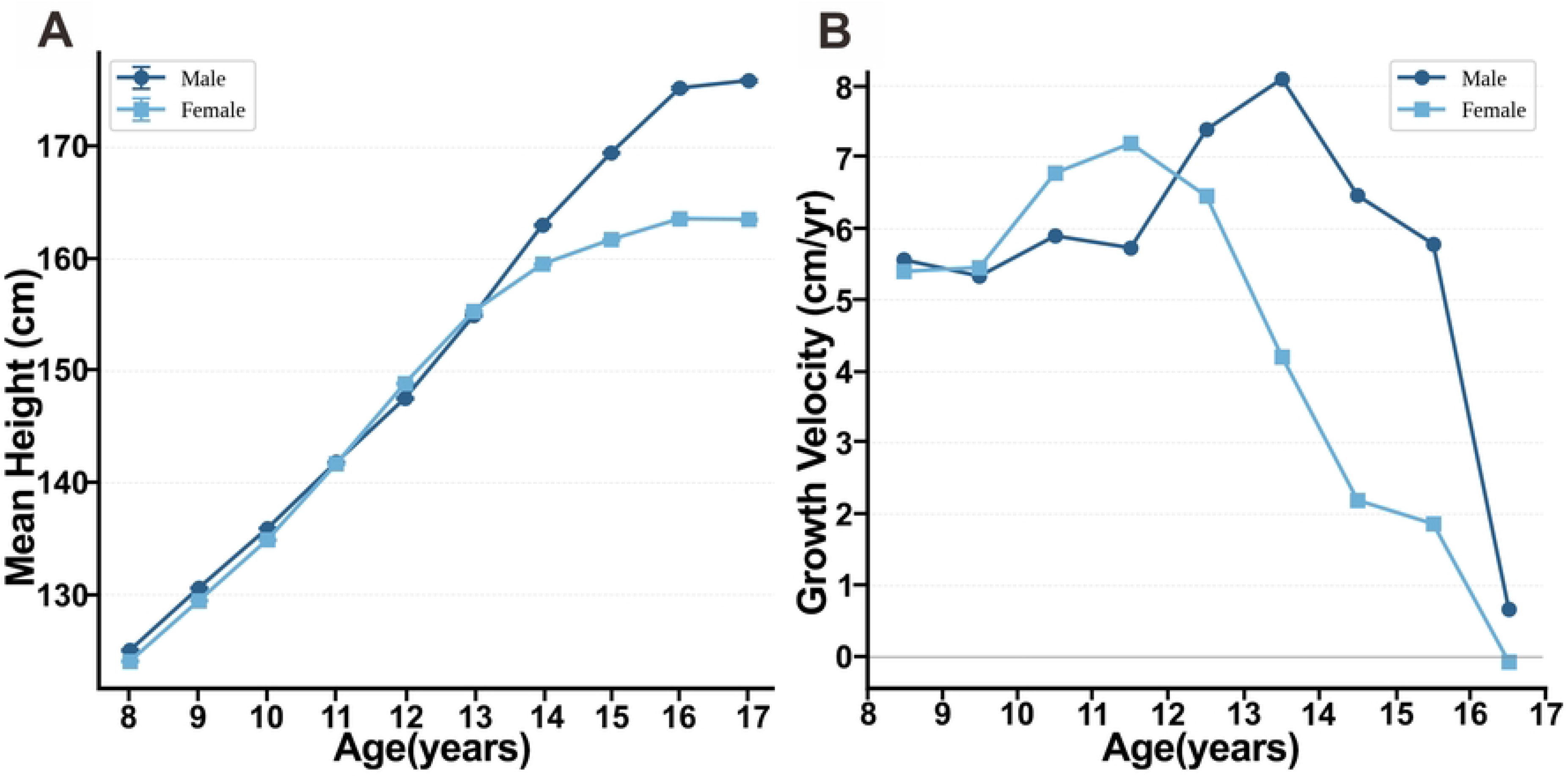
Age- and sex-specific distributions of resting heart rate and BMI among adolescents. Note: (A) Age-group mean resting heart rate (RHR): bars denote average RHR, error bars show 95% mean confidence intervals. (B) Floating boxplots present interquartile range (IQR, Q1-Q3). Thick horizontal lines stand for medians, red diamonds for means; whiskers cover values within 1.5×IQR. Welch’s t-test: t = −14.663, p = 0.0001 ***; Cohen’s d = −0.081 (negligible effect). (C) Violin plots illustrate annual BMI distributions of boys and girls aged 8-17. Outer contours show BMI kernel density, dashed lines IQR, solid white lines medians. Boys have higher median BMI and more high-BMI outliers across nearly all ages. Such gender gaps grow wider in early and mid-adolescence, supporting a significant age-sex interaction for BMI trends.

### 3.5 Multifactorial Interactions

This multidimensional analysis identified prominent modifying effects of age and sex on the correlations among BMI, UCVA and resting heart rate (RHR), revealing complex multifactorial interactions across adolescent developmental stages. A significant age-sex interaction was detected for BMI via two-way ANOVA (F=58.83, p=1.73×10^^−14^) (Figure. S2). Boys consistently exhibited higher median BMI and a larger proportion of high-BMI outliers than girls, and the gender gap in BMI widened notably from early to mid-adolescence. A substantial BMI-age interaction also existed for visual acuity (F=14.74, p<0.001): the protective association between higher BMI and better log MAR UCVA was strongest in children aged 8-11 years, weakened among 12-16-year-olds, and reversed to a negative correlation in 17-year-olds. Moreover, the age-related worsening of eyesight and expanded individual variability in visual performance coincided with dramatic BMI shifts during puberty, confirming age and sex jointly modulate the magnitude and direction of the BMI-vision association. For resting heart rate, two-way ANOVA verified a significant age-sex interaction (F=12.05, p<0.001), meaning the annual descending rate of RHR differed slightly between males and females. Though age and sex were statistically meaningful predictors of RHR, they only explained a tiny fraction of heart rate variance (R^^2^=0.033), indicating autonomic regulation of resting heart rate is governed mainly by unmeasured physiological factors such as cardiorespiratory fitness. Collectively, age and sex act as core moderators shaping the developmental trajectories of BMI, vision and heart rate, as well as the pairwise relationships between these physiological indicators. Shared behavioral and biological mechanisms including near-work load, outdoor activity duration, sleep and screen-use patterns, and endocrine changes underpin these interactive patterns. The evident multifactorial interactions highlight the necessity of developmental-stage and gender-stratified integrated health interventions for adolescents, instead of isolated single-risk prevention strategies for obesity, myopia or cardiovascular abnormalities.

## 4. Discussion

This study provides an integrated analysis of health screening data from 130,832 examination records of adolescents aged 8-17 years, revealing a complex, interconnected relationship between BMI, UCVA, and resting heart rate, with significant variation by sex and age. Boys had higher BMI and greater overweight/obesity prevalence, while girls showed worse UCVA across all ages. These findings highlight that adolescent health outcomes are shaped by the interplay of biological, behavioral, and developmental factors, suggesting that single-issue intervention approaches may have limited effectiveness.

The research reveals a pronounced sexual dimorphism in the relationship between BMI and visual acuity. Male adolescents not only demonstrate superior mean UCVA but also greater interindividual variability compared to females. This divergence may be attributed to the influence of pubertal sex hormone levels and sex-differentiated patterns of visual behavior ^[30–32]^. Moreover, the association between BMI and visual acuity peaks during early adolescence (ages 12-16), implicating this period as a critical window. The underlying mechanisms may involve the synergistic effects of growth hormone surges and insulin sensitivity changes characteristic of this developmental stage, with elevated BMI potentially exacerbating ocular developmental processes through the induction of inflammatory and oxidative stress responses ^[33]^.

A pronounced disparity in obesity rates was observed, with male adolescents exhibiting significantly higher rates than females, a difference stemming from the interplay of physiological characteristics and sex-based behavioral roles^[34]^ . Clinically, this is highly significant. Since obesity is an established independent risk factor for cardiovascular disease^[19, 20]^ and male obese adolescents demonstrate a greater propensity for metabolic syndrome ^[35]^, this demographic must be recognized as a high-risk group. The pathophysiological basis for this risk is largely underpinned by obesity-related autonomic nervous system imbalance, manifesting as sympathetic overactivity and diminished vagal modulation^[36, 37]^. This autonomic dysfunction is directly reflected in an accelerated resting heart rate. In our data, the association between BMI and resting heart rate was statistically significant but modest in magnitude. Nevertheless, the monitoring of resting heart rate offers a dual utility: it facilitates the early detection of impaired cardiac autonomic regulation and provides a straightforward, practical indicator for gauging long-term cardiovascular risk in this population.

Using a large-scale, multi-year physical examination dataset, this study simultaneously examined the associations of BMI with both visual acuity and resting heart rate in adolescents. The use of multivariable regression models allowed us to adjust for key confounders and explore effect modification by sex and age. The observed associations, while modest in magnitude, are consistent with previous reports ^[15, 16, 23]^ and suggest that effective adolescent obesity management may confer ancillary benefits for visual and cardiovascular health. Family- and school-based interventions that promote healthy lifestyle habits-such as regular breakfast consumption, adequate physical activity, reduced screen time, and optimized sleep patterns-have demonstrated efficacy in improving BMI and related health outcomes ^[38–41]^. However, the extent to which such interventions can simultaneously improve visual acuity and heart rate regulation remains to be demonstrated in prospective interventional studies.

However, this study has several limitations. Notably, its reliance on physical examination data means that certain variables-such as behavioral factors, academic stress, and environmental influences-may have been measured with limited precision or could be subject to recall bias. Furthermore, it was not possible to account for all potential confounding factors (e.g., genetic predispositions and detailed dietary intake), and the generalizability of the findings may be constrained by the sampling frame (e.g., the inclusion of specific regions or schools). Future research should aim to incorporate more precise measurements of these factors to facilitate a more comprehensive, multi-level analysis.

## 5. Conclusions

In 130,832 health-examination records from adolescents aged 8-17 years, BMI showed small, age- and sex-dependent associations with UCVA and RHR. Boys had higher BMI distributions and a greater burden of overweight and obesity, whereas reduced UCVA was more common among girls. After covariate adjustment, higher BMI was associated with slightly lower UCVA log MAR, with a stronger inverse coefficient in girls than boys. The association was largest at ages 8-11 years, attenuated at ages 12-16 years, and reversed at age 17; the late-adolescent finding should be interpreted cautiously because few records were available at ages 16-17. Higher BMI was also associated with a small increase in RHR (0.26 beats/min per kg/m²), while BMI, age and sex are also correlated with resting heart rate to a certain extent.

These findings support integrated but differentiated adolescent health screening. Weight-management resources should give particular attention to boys and to the period when high-BMI upper tails begin to widen, while visual screening should remain especially attentive to girls and to the progressive age-related decline in UCVA. RHR can be included as a low-burden cardiovascular measure, but it should be interpreted with age, sex, measurement conditions, fitness, and clinical context. The inverse BMI-UCVA association must not be interpreted as evidence that higher BMI improves or protects vision.

Because the study was observational and lacked detailed body-composition, behavioral, cardiovascular, and ocular measurements, causal inference is not possible. Future prospective studies should incorporate body-fat percentage, waist-to-height ratio, pubertal stage, blood pressure, cardiorespiratory fitness, objective physical activity and outdoor exposure, near-work and screen-time measures, cycloplegic refraction, and ocular axial length. Such data are needed to determine whether the reported associations reflect shared developmental or environmental determinants and to test possible mediating pathways.

## 7. Data availability

The de–identified minimal dataset supporting the conclusions of this study is available as cleaned_data.csv file within the Supporting Information. All relevant metadata are included within the manuscript. No additional restrictions apply to the data.

## Data Availability

Data availability The complete, de-identified dataset (including all physical examination indicators for all participants) supporting the findings of this study is available as supplementary information files (cleaned_data.csv) accompanying this article.

## Acknowledgements

We thank all school health–screening staff, adolescents and their guardians who participated in this study. We also thank the hospital research administrative team for administrative support during this work.

## Supporting Information

Figure S1. Cross-sectional stature and derived annual growth rates among children and adolescents aged 8 to 17 years stratified by sex (Supplementary.docx).

Figure S2. Age- and sex-specific mean resting heart rate from ages 8 to 17 years (Supplementary.docx).

